# Ex Vivo Immune Profiling Defines a Continuous Functional Immune Axis and a Sepsis-Enriched Low-Response State in Critical Illness

**DOI:** 10.64898/2026.04.28.26351971

**Authors:** Ruth-Ann Brown, Anthony S Bonavia

**Author notes:** Corresponding author:* Anthony Bonavia, MD.

## Abstract

**Background:** Immune dysfunction in sepsis and critical illness is biologically heterogeneous, yet available stratification frameworks leave many patients unclassified. We hypothesized that ex vivo cytokine-induction responses would define a continuous axis of functional immune responsiveness and identify a low-response state enriched in sepsis.

**Methods:** In this prospective observational study, 39 critically ill adults enrolled within 48 hours of ICU admission and 6 healthy controls underwent standardized whole-blood stimulation with lipopolysaccharide, anti-CD3/anti-CD28 antibodies, and PMA/ionomycin, with selected wells additionally supplemented with interleukin-7 or granulocyte-macrophage colony-stimulating factor. Interleukin-6, tumor necrosis factor, and interferon-gamma responses were quantified and referenced to subject-specific unstimulated baselines. A patient-anchored primary feature matrix was used to derive a continuous immune axis by principal component analysis, and a cross-validated 5-feature *MiniResponder* score was developed as a portable summary measure.

**Results:** Among critically ill patients, induced cytokine responses organized along a dominant continuous axis of functional immune responsiveness; the first principal component explained 53.3% of between-patient variance. *MiniResponder* captured this axis and showed a lower-shifted distribution in sepsis. Using a control-referenced threshold defined by the 10th percentile of the healthy-control distribution, 19 of 39 patients (48.7%) were classified as low-response, including 15 of 21 patients with sepsis (71.4%) and 4 of 18 critically ill patients without sepsis (22.2%) (odds ratio 8.75, Fisher exact *P=*0.004). In exploratory analyses, lower *MiniResponder* scores were associated with greater unadjusted improvement in Sequential Organ Failure Assessment score from day 1 to days 3-9 (rho=-0.33; P=0.046), but this association attenuated after adjustment for baseline SOFA score (beta=-0.10; 95% CI-0.36 to 0.27).

**Conclusions:** Ex vivo immune profiling identified a continuous patient-anchored axis of functional immune responsiveness in critical illness that can be summarized by a compact 5-feature score. A control-referenced low-response state was enriched in sepsis. This framework may complement existing biomarker-based stratification approaches and support future enrichment strategies in sepsis trials.

## Introduction

Sepsis remains a leading cause of death and long-term morbidity worldwide, accounting for an estimated 11 million deaths annually despite advances in supportive care. A central challenge is its marked biological heterogeneity: patients meeting identical clinical criteria may exhibit divergent immune states ranging from hyperinflammation to profound immunosuppression (1, 2). This heterogeneity likely underlies repeated failures of immunotherapy trials in unselected populations, where treatment effects in responsive subgroups are diluted (3, 4).

Current understanding frames sepsis immune dysregulation as a dynamic and overlapping spectrum rather than a linear trajectory (5). Transcriptomic endotyping approaches, including SRS, MARS, and related frameworks, have identified biologically coherent subphenotypes associated with differential outcomes and treatment responses (6–8). For example, re-analysis of the VANISH trial demonstrated increased mortality with hydrocortisone in SRS2 but not SRS1 patients (9). However, transcriptomic approaches remain impractical for routine clinical use due to cost, complexity, and turnaround time (10).

Protein biomarker strategies offer a more feasible alternative. Two clinically relevant states include macrophage activation-like syndrome, associated with hyperinflammation and elevated ferritin, and sepsis-induced immunoparalysis, defined by low mHLA-DR expression (11, 12). These states have corresponding therapeutic targets, including anakinra and IFN-gamma (13, 14). The PROVIDE trial provided early proof-of-concept for biomarker-guided immunotherapy (11), and the recent ImmunoSep trial confirmed that such strategies can improve organ dysfunction in selected patients (15). However, most patients fall into an “unclassified” intermediate state not addressed by current frameworks.

mHLA-DR, while widely used, reflects only one dimension of immune function and does not capture broader inducible cytokine responses across innate and adaptive pathways (16–18). Prior work shows limited correlation between mHLA-DR and ex vivo cytokine production, suggesting complementary information in functional assays (19, 20). Standardized ex vivo stimulation (e.g., lipopolysaccharide (LPS), anti-CD3/CD28, and phorbol 12-myristate 13-acetate (PMA) stimulation) provides an integrated assessment of immune responsiveness.

Using a rapid microfluidic platform, we previously demonstrated that induced cytokine responses are reproducible, dynamic, and associated with organ dysfunction in critical illness (20–22). These findings motivate evaluation of whether integrated cytokine responses define a coherent functional immune axis that may extend beyond existing biomarker classifications.

In this study, we apply a compact ex vivo immune profiling panel in critically ill adults and derive a patient-anchored functional immune axis using principal component analysis and a cross-validated *MiniResponder* score. We show that immune responsiveness is continuous, that a low-response state is enriched in sepsis, and that this functional framework may help characterize patients not captured by current biomarker-based stratification strategies. Throughout this manuscript, we treat *MiniResponder* as a portable summary of a continuous patient-anchored immune axis, and the low-response state as a control-referenced enrichment definition within that continuum.

## Methods

### Study Design and Population

We conducted a prospective observational study of critically ill adults admitted to a tertiary academic intensive care unit (ICU) (IRB #15328). Eligible patients were enrolled within 48 hours of ICU admission and did not have major immune-modifying conditions, including recent cytotoxic chemotherapy, hematologic malignancy, or chronic immunosuppression. Sepsis was defined according to Sepsis-3 criteria (23). A parallel cohort of healthy volunteers was studied using identical procedures to provide a reference distribution for immune responsiveness (IRB #27044). The study was approved by the Penn State Institutional Review Board.

### Functional Cytokine Response Profiling

Venous whole blood was diluted 1:10 in cell-culture media and incubated for 18 to 22 hours under standardized stimulation conditions representing myeloid and T/NK activation pathways, including media control, LPS, anti-CD3/anti-CD28 antibodies, and PMA/ionomycin. Selected wells additionally included IL-7 or GM-CSF to assess whether cytokine co-stimulation augmented latent response capacity. Cytokine responses (IL-6, TNF, and IFN-gamma) were measured using the Ella multiplex immunoassay system (Bio-Techne, Minneapolis, Minnesota). Experimental procedures followed previously described protocols (19, 21, 22).

To focus on functional immune responsiveness rather than resting inflammatory tone, stimulated cytokine values were referenced to each subject’s unstimulated baseline. These induced-response features formed the basis of the downstream multivariable analyses.

### Derivation of the Patient-Anchored Immune Axis

The primary analysis used an a priori matched-stimulus feature set derived from induced cytokine responses under LPS, CD3/28, and PMA stimulation conditions, with or without the matched adjuvant. Unsupervised analyses were performed using critically ill patients only to avoid conflating health-disease separation with heterogeneity among critically ill patients. Healthy controls were projected into the resulting feature space afterward using preprocessing parameters derived from patients.

Features with high missingness were excluded and remaining missing values were imputed and standardized using patient-only data. The dominant immune-response axis was defined as the first principal component of the patient-only matched-stimulus matrix. Axis orientation was fixed so that higher values corresponded to greater overall inducible responsiveness. Prespecified sensitivity analyses addressing feature-set selection and axis stability are described in the Supplementary Methods and Supplementary Figure 1.

### *MiniResponder* Score and Low-Response State

To obtain a portable continuous summary of inducible immune responsiveness, we derived a 5-feature *MiniResponder* score in a cross-validated manner designed to avoid overfitting. For each held-out patient, score derivation was repeated using training patients only, such that the held-out patient received an out-of-fold score. A final 5-feature panel was then fit for descriptive analyses and figure generation. Sepsis status, baseline clinical variables, and SOFA outcomes were not used to construct the latent immune axis or derive the *MiniResponder* score.

For exploratory translational analyses, a control-referenced low-response state was defined using the healthy-control *MiniResponder* distribution. Patients below the 10th percentile of controls were classified as low-response. Patients above this threshold were not treated as a biologically homogeneous comparator subtype, but rather as the remainder of a continuous functional spectrum. Alternate threshold definitions are reported in the Supplementary Methods.

### Exploratory SOFA anchoring analyses

Cross-sectional clinical associations with *MiniResponder* were assessed among patients only. Continuous variables were summarized using Spearman correlations. Binary variables were explored by comparing *MiniResponder* distributions with Mann-Whitney tests and, when applicable, by comparing low-response-state frequencies with Fisher exact tests.

Serial SOFA scores were available every 48 h between days 1-15 of study inclusion. To provide exploratory clinical anchoring, we examined associations between *MiniResponder* and adapted SOFA improvement endpoints aligned conceptually with the organ-dysfunction framework used in recent precision-immunotherapy sepsis studies (15). The primary SOFA endpoint was day 1 SOFA minus the mean SOFA across days 3, 5, 7, and 9, with larger values indicating greater improvement in organ dysfunction. Secondary SOFA endpoint definitions are provided in the Supplementary Methods. Because this was an observational pilot study rather than a treatment trial, SOFA analyses were interpreted as exploratory associations rather than efficacy tests.

To assess whether associations between *MiniResponder* and exploratory SOFA anchoring analyses were independent of baseline illness severity, we fit baseline-adjusted regression models including day 1 SOFA and *MiniResponder* as predictors. Confidence intervals for the *MiniResponder* term were estimated using resampling methods described in the Supplementary Methods.

### Statistical Analysis

All analyses were performed in Python using standard scientific-computing libraries. Continuous variables are reported as medians with interquartile ranges or means with standard deviations as appropriate, and categorical variables as counts and proportions. Given the pilot and hypothesis-generating nature of the study, emphasis was placed on effect sizes, rank concordance, and resampling-based precision rather than on dichotomous significance testing alone.

This study was designed as an exploratory precision-oriented pilot intended to assess whether a coherent inducible immune-response axis could be recovered from a compact ex vivo cytokine panel and whether that axis could support future enrichment strategies. Formal power calculations were not used to determine sample size.

Throughout this manuscript, we use functional immune responsiveness to describe the underlying biologic construct, *MiniResponder* to describe its portable 5-feature summary score, and low-response state to describe the control-referenced enrichment definition applied to that continuous axis.

## Results

### Cohort and Cytokine Data

We obtained complete or near-complete cytokine-induction profiles for 45 individuals: 39 critically ill patients and 6 healthy volunteers. Among patients, 21 met sepsis diagnostic criteria and 18 were critically ill without sepsis (**Table 1**). The non-baseline analytic matrix contained 24 induced-response features spanning IL-6, TNF, and IFN-gamma across matched stimulant and adjuvant conditions. The primary matched-stimulus matrix contained 18 features. Feature missingness was low; among patients, all primary features were missing in 5.1% or fewer subjects and none exceeded the prespecified 30% exclusion threshold.

**Table 1:** Demographic and Clinical Characteristics of Critically Ill Patients, Overall and by Sepsis Status.

| Characteristic | Overall<br>(N=39) | Critically ill non-sepsis<br>(N=18) | Sepsis<br>(N=21) | *P* value |
| --- | --- | --- | --- | --- |
| **Demographic characteristics** |  |  |  |  |
| Age, y | 69 [53-76] | 72 [58-77] | 64 [51-76] | 0.554 |
| Female sex, No./total No. (%) | 17/39 (43.6%) | 7/18 (38.9%) | 10/21 (47.6%) | 0.748 |
| White race, No./total No. (%) | 33/39 (84.6%) | 14/18 (77.8%) | 19/21 (90.5%) | 0.387 |
| Body mass index | 29.4 [25.0-35.6] | 28.6 [25.1-36.2] | 31.3 [24.2-35.0] | 0.833 |
| **Severity of illness and support** |  |  |  |  |
| APACHE II score | 21 [18-26] | 22 [17-24] | 21 [19-28] | 0.381 |
| SOFA score at enrollment | 8 [5-10] | 8 [4-9] | 9 [6-12] | 0.090 |
| Blood lactate | 4.0 [2.4-5.2] | 2.4 [1.8-4.3] | 4.3 [3.0-5.2] | 0.202 |
| Mechanical ventilation, No./total No. (%) | 26/38 (68.4%) | 16/18 (88.9%) | 10/20 (50.0%) | 0.015 |
| Vasopressor infusion, No./total No. (%) | 24/38 (63.2%) | 8/18 (44.4%) | 16/20 (80.0%) | 0.042 |
| Septic shock, No./total No. (%) | 15/39 (38.5%) | 0/18 (0.0%) | 15/21 (71.4%) | <0.001 |
| **Laboratory values** |  |  |  |  |
| White blood cell count | 14.9 [9.0-19.8] | 12.7 [8.0-16.5] | 18.5 [11.0-27.0] | 0.047 |
| Absolute neutrophil count | 12.0 [6.4-18.1] | 9.8 [5.9-14.2] | 16.9 [8.3-24.6] | 0.094 |
| Absolute lymphocyte count | 0.81 [0.45-1.12] | 0.97 [0.83-1.24] | 0.46 [0.30-0.91] | 0.014 |
| Platelet count | 180 [108-263] | 183 [143-257] | 152 [95-302] | 0.535 |
| Creatinine | 1.32 [0.98-1.73] | 1.25 [0.80-1.52] | 1.44 [1.05-2.34] | 0.390 |
| Total bilirubin | 0.8 [0.4-1.6] | 0.4 [0.4-1.4] | 0.8 [0.5-1.6] | 0.284 |
| **Clinical outcomes** |  |  |  |  |
| 28-day ICU-free days | 22 [17-26] | 18 [8-26] | 23 [19-27] | 0.152 |
| Hospital length of stay, d | 9 [6-16] | 7 [5-12] | 10 [7-20] | 0.124 |
| In-hospital mortality, No./total No. (%) | 4/39 (10.3%) | 3/18 (16.7%) | 1/21 (4.8%) | 0.318 |
| **Primary immune analysis** |  |  |  |  |
| MiniResponder score | 0.34 [-0.97-1.77] | 1.77 [0.41-3.23] | -0.37 [-2.46-0.34] | <0.001 |
| Control-referenced low-response state, No./total No. (%) | 19/39 (48.7%) | 4/18 (22.2%) | 15/21 (71.4%) | 0.004 |

*Values are reported as median [IQR] unless otherwise indicated.*
Abbreviations: APACHE II, Acute Physiology and Chronic Health Evaluation II; ICU, intensive care unit; IQR, interquartile range; SOFA, Sequential Organ Failure Assessment. Percentages were calculated using nonmissing denominators. P values compare critically ill non-sepsis vs sepsis groups using Mann-Whitney U tests for continuous variables and Fisher exact tests for categorical variables. Baseline SOFA was taken from the day-1 SOFA dataset used in the exploratory SOFA analysis. *MiniResponder* and the control-referenced low-response state were derived from the primary matched-stimulus patient-anchored immune analysis.

### Inducible Cytokine Responses Define a Dominant Immune-Response Axis

Across critically ill patients, induced cytokine responses organized along a single dominant axis of functional immune responsiveness (**Figure 1**). In the primary matched-stimulus matrix, the first principal component explained 53.3% of between-patient variance. Bootstrap resampling yielded a median PC1 variance explained of 55.0%, with a 95% CI from 39.8% to 69.0%, supporting a reproducible one-dimensional structure.

**Figure 1.**
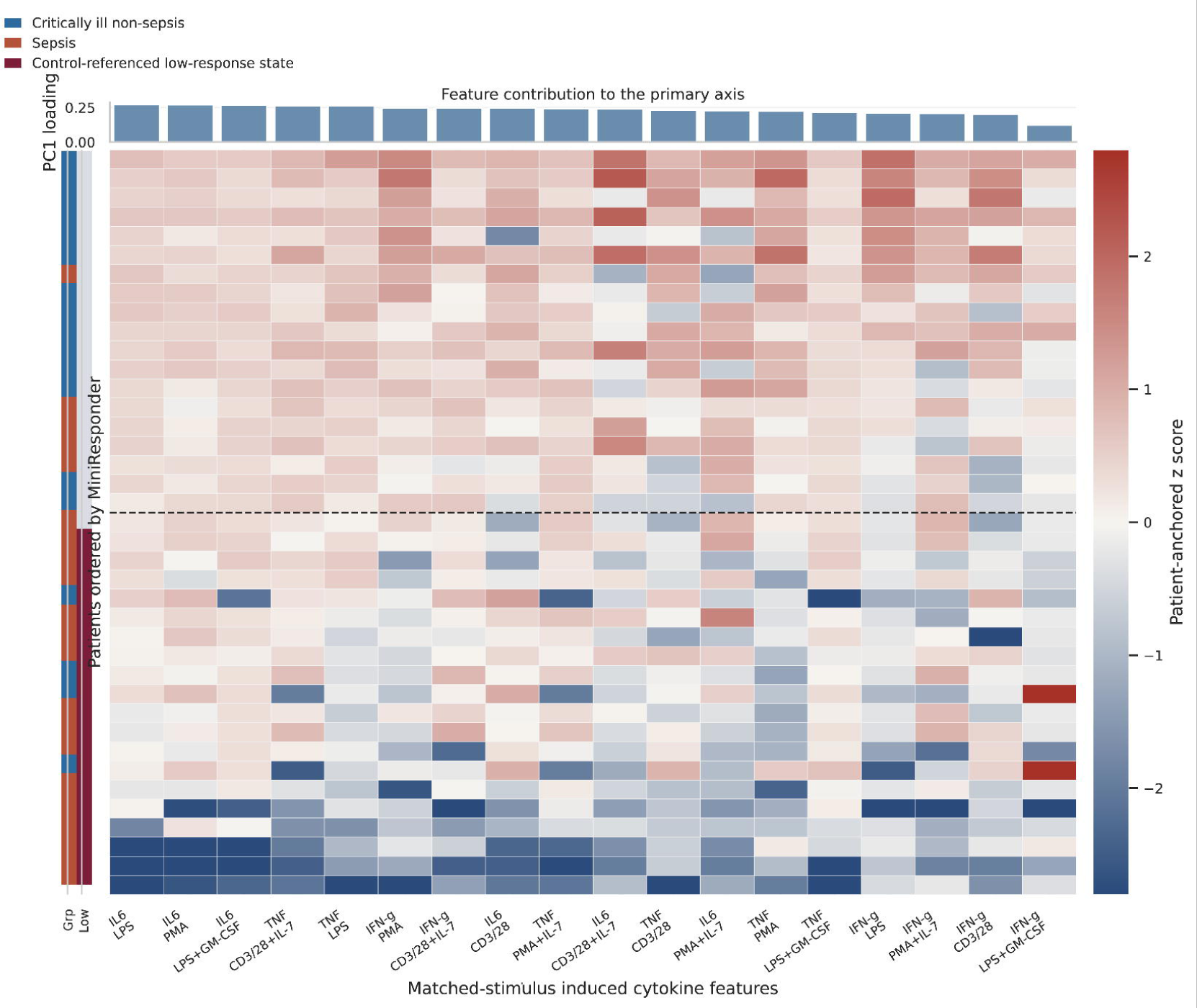
Patient immune-response heatmap ordered along the inducible axis. Heatmap of patient-only standardized induced cytokine responses across the 18 matched-stimulus features used in the primary analysis. Rows represent individual patients ordered from highest to lowest *MiniResponder* score, so that adjacent rows reflect nearby positions along the dominant functional immune axis. Left-side annotations show clinical group and the control-referenced low-response state. Columns are ordered by absolute loading on the first patient-derived principal component, and the bar plot above the heatmap shows the corresponding feature loadings. Warmer colors indicate relatively stronger induced responses and cooler colors indicate relatively weaker induced responses after patient-anchored preprocessing. Rows: n=39 patients. Columns: n=18 matched-stimulus features. Dashed line marks the low-response subgroup (n=19).

Feature loadings indicated that this axis was driven principally by coordinated induced responses in IL-6, TNF, and IFN-gamma under LPS-, PMA-, and CD3/28-based stimulation, rather than by any single cytokine or condition. The strongest contributors included IL-6 responses to LPS and PMA, TNF responses to LPS and CD3/28 + IL-7, and IFN-gamma responses to PMA and CD3/28 + IL-7. Heatmap visualization showed a graded rather than sharply segmented organization of patients along this functional axis (**Figure 1**).

### *MiniResponder* Captures the Dominant Functional Axis and Identifies a Sepsis-Enriched Low-Response State

The nested cross-validated *MiniResponder* workflow identified a stable 5-feature panel consisting of IL6(cytokine)-LPS(stimulant)-none(adjuvant), IFN-gamma-PMA-none, TNF-LPS-none, IFN-gamma-LPS-none, and TNF-PMA-none; four of these five features were selected in all 39 leave-one-out folds, and the fifth was selected in 38 of 39 folds. *MiniResponder* values varied continuously across subjects, with healthy controls and critically ill patients without sepsis occupying overlapping higher portions of the score distribution and sepsis patients showing a broader, lower-shifted distribution (**Figure 2A**).

**Figure 2.**
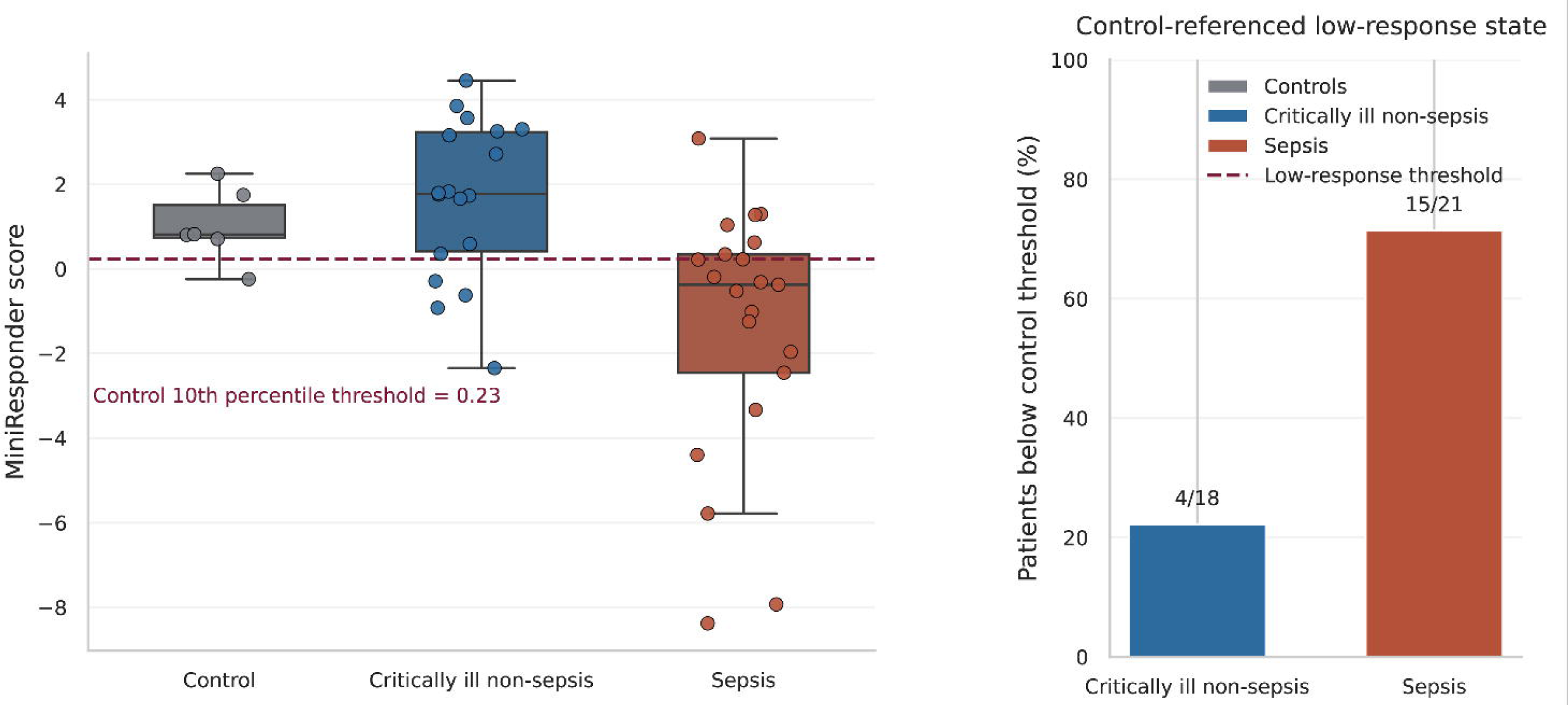
*MiniResponder* distribution and low-response enrichment. Panel A shows the distribution of *MiniResponder* scores in controls, critically ill non-sepsis patients, and sepsis patients. Low-response state was defined relative to the control distribution and used as an enrichment-style subgroup rather than as a hard biological class. The dashed horizontal line marks the primary low-response threshold, defined as the 10th percentile of the healthy-control distribution. Panel B shows the proportion of patients in each clinical group falling below that control-referenced threshold. *MiniResponder* is presented primarily as a continuous functional score while also illustrating how a threshold-defined enrichment subgroup can be derived without outcome tuning.

Using the prespecified control-referenced enrichment rule, the 10th percentile of the healthy-control *MiniResponder* distribution corresponded to a threshold of 0.23. Nineteen of 39 patients (48.7%) fell below this threshold and were designated as belonging to the low-response state. This enrichment state was strongly associated with sepsis: 15 of 21 sepsis patients (71.4%) versus 4 of 18 critically ill patients without sepsis (22.2%) met the low-response criterion (odds ratio, 8.75; Fisher exact *P* = 0.0036) (**Figure 2B**). These findings support *MiniResponder* as both a continuous descriptor of inducible immune responsiveness and a control-anchored enrichment variable for future translational studies.

### The Immune Axis Is Biologically Coherent and Robust to Feature-Set Variation

*MiniResponder* was positively associated with absolute lymphocyte count (Spearman rho = 0.44; *P* = 0.005) and absolute monocyte count (rho = 0.47; *P* = 0.002) but showed only weak associations with total white blood cell count, neutrophil count, baseline SOFA score, APACHE II score, or organ-dysfunction-free days. These effect sizes suggest that the axis captures functional immune responsiveness that is related to, but not reducible to, conventional hematologic measurements. Lower *MiniResponder* scores were observed among patients with shock (mean difference for shock vs no shock, −2.23; Mann-Whitney *P =* 0.018), whereas associations with mortality and secondary infection were directionally heterogeneous and underpowered for firm inference.

Sensitivity analyses showed that the full 24-feature non-baseline axis was almost identical to the primary matched-stimulus axis (Spearman rho = 0.99), indicating that exclusion of adjuvant-only wells did not materially alter patient rank ordering. Removal of all adjuvant conditions produced a more compressed but still highly concordant axis (rho = 0.92), whereas the baseline-only matrix showed substantially weaker concordance with the primary axis (rho = 0.62). Dynamic range along the axis decreased from 10.4 units in the primary matched-stimulus analysis to 6.8 units in the no-adjuvant analysis, consistent with cytokine adjuvants amplifying contrast along an existing functional axis rather than redefining patient ordering.

### Exploratory SOFA anchoring analyses

Serial SOFA data were available for all 39 patients on day 1, with increasing missingness at later time points due to the critically ill nature of the patient population (missing values: day 3, n = 1; day 5, n = 4; day 7, n = 15; day 9, n = 20; day 15, n = 28). In unadjusted analyses, lower *MiniResponder* scores were associated with greater apparent improvement in organ dysfunction. For the SOFA endpoint, defined as day 1 SOFA minus the mean SOFA across days 3, 5, 7, and 9, *MiniResponder* correlated inversely with SOFA improvement (Spearman rho = −0.33; *P* = 0.046). Patients in the control-referenced low-response state showed greater median improvement than the remaining patients (6.0 vs 3.5 SOFA points; Mann-Whitney *P* = 0.032). Similar directional results were observed for the day 15 endpoint and under LOCF sensitivity analyses (**Figure 3**).

**Figure 3.**
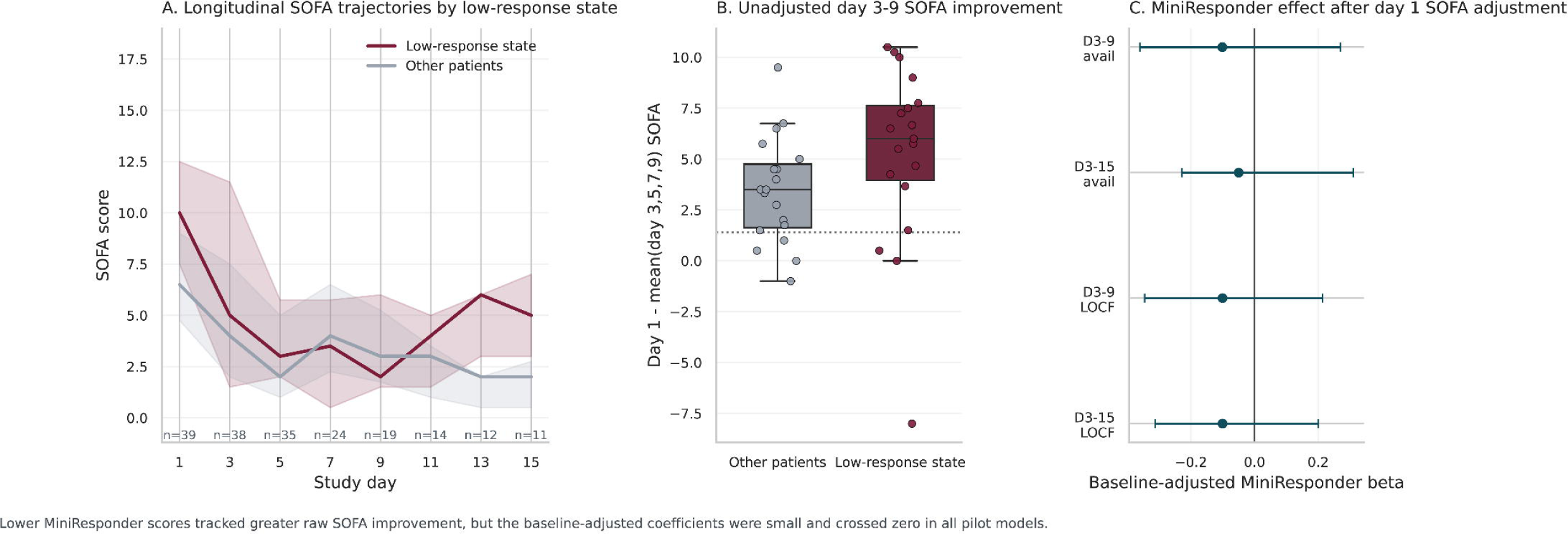
Exploratory SOFA anchoring analyses. Panel A shows SOFA trajectories from day 1 through day 15, summarized as median values with interquartile ribbons for patients in the control-referenced low-response state versus all other patients. Labels beneath the x-axis show the number of patients with available SOFA values at each day. Panel B shows the unadjusted primary pilot SOFA endpoint: day 1 SOFA minus the mean SOFA across days 3, 5, 7, and 9, with larger values indicating greater improvement in organ dysfunction. The dotted line indicates the 1.4-point threshold adopted from the ImmunoSep trial. Panel C shows baseline-adjusted *MiniResponder* coefficients for day 9 and day 15 SOFA-improvement endpoints under available-case and LOCF sensitivity definitions. Lower *MiniResponder* scores were associated with greater raw SOFA improvement, but that this relationship attenuated after accounting for baseline day 1 SOFA.

However, *MiniResponder* was also inversely associated with baseline day 1 SOFA score (Spearman rho = −0.40; *P* = 0.011), indicating that lower-response patients entered follow-up with greater baseline organ dysfunction and therefore more opportunity for raw improvement. After adjustment for day 1 SOFA, the association between *MiniResponder* and the primary day 3-to-day 9 SOFA-improvement endpoint attenuated substantially (beta = −0.10; bootstrap 95% CI, −0.36 to 0.27). Baseline-adjusted estimates for day 15 and LOCF-derived endpoints were similarly small, and all confidence intervals crossed zero. Binary achievement of a 1.4-point or greater SOFA-improvement threshold was common and was not meaningfully discriminated by *MiniResponder* or by the low-response enrichment state. Taken together, these pilot analyses suggest that lower *MiniResponder* scores track greater raw SOFA change, but that much of this relationship may reflect higher baseline illness severity rather than an independent association with subsequent organ-dysfunction recovery.

## Discussion

This pilot study demonstrates that ex vivo inducible cytokine responses across multiple stimulation pathways organize along a single, dominant, and continuous axis of functional immune responsiveness in critical illness. This axis is reproducible, primarily reflects inducible rather than baseline immune activity, and can be summarized by a compact *MiniResponder* score. A control-referenced low-response state was strongly enriched among patients with sepsis, while associations with longitudinal SOFA improvement were largely explained by baseline illness severity. The conceptual model advanced by these data is a continuous patient-anchored axis of functional immune responsiveness, with threshold-based enrichment superimposed for translational use.

A central implication of these findings is that immune dysfunction in critical illness may be more appropriately conceptualized as a continuous quantitative trait rather than a set of discrete categories. This contrasts with prevailing classification approaches, including ferritin/mHLA-DR-based stratification (11, 12), transcriptomic endotypes (6, 7, 24), and clinical phenotypes (25), which assign patients to fixed groups. The limited concordance observed across these frameworks suggests that each captures only a subset of a shared underlying biology (24). A continuous representation of immune responsiveness may therefore better support threshold-based enrichment and modeling of heterogeneity of treatment effect (10).

The ImmunoSep randomized clinical trial provides an important clinical context for interpreting these findings (15). That study established that biomarker-guided immunotherapy can improve organ dysfunction in selected patients with sepsis, using ferritin and mHLA-DR to define treatment groups. However, more than half of screened patients did not meet criteria for either enrichment state, and were excluded. This “unclassified” population represents a major gap in current precision immunotherapy strategies. The *MiniResponder* axis, derived from integrated functional responses across innate and adaptive pathways, may help describe heterogeneity within this group by identifying biologically meaningful heterogeneity within this large intermediate group. In contrast to static, single-timepoint thresholds, a continuous functional score could support adaptive enrichment, stratified trial designs, or biomarker–treatment interaction analyses.

Our findings also reinforce the partial independence of functional immune responsiveness from mHLA-DR. Prior studies have shown weak correlation between HLA-DR expression and stimulated cytokine production (19), and our results are consistent with this dissociation. By incorporating both myeloid and lymphocyte-driven responses, the *MiniResponder* framework captures a broader representation of immune function, including adaptive immune deficits that may not be detected by mHLA-DR alone.

The use of a control-referenced threshold provides a biologically grounded approach to defining an enrichment state, anchored to deviation from healthy immune function rather than cohort-specific distributions (26). This strategy was robust across sensitivity analyses and may offer practical advantages for reproducibility across study populations.

Inclusion of cytokine adjuvants (IL-7, GM-CSF) increased dynamic range without altering patient rank ordering, suggesting that these conditions amplify latent immune capacity rather than introduce additional dimensions of variation. This supports their potential utility in pharmacodynamic monitoring rather than primary phenotyping.

Interpretation of the exploratory SOFA anchoring analyses requires caution. Although lower *MiniResponder* scores were associated with greater unadjusted improvement in organ dysfunction, this relationship was largely attenuated after adjustment for baseline severity.

Patients with lower scores had higher initial SOFA values and therefore greater opportunity for numerical improvement. This underscores a broader limitation of SOFA trajectory analyses and highlights the importance of severity-adjusted or threshold-based endpoints, such as the ≥1.4-point improvement used in ImmunoSep (15).

This study has several limitations. The sample size is modest and limits power for clinical outcome associations. The healthy control cohort is small, which may affect precision of control-referenced thresholds. The study was conducted at a single center, and external validation in larger, multicenter cohorts will be required. In addition, the assay currently requires timely processing and overnight incubation, which may limit immediate clinical applicability. Finally, all clinical associations should be considered hypothesis-generating given the observational design.

Despite these limitations, this work provides a practical framework for functional immune profiling in critical illness. Its principal translational relevance is that it may characterize patients who remain unclassified by existing biomarker strategies. Integration of functional inducibility with established markers such as mHLA-DR may ultimately provide a more comprehensive immune phenotype, enabling more precise selection of patients for immunomodulatory therapies.

In conclusion, inducible cytokine responses define a continuous functional immune axis in critical illness that can be captured by a compact, reproducible score. This framework complements existing biomarker-based approaches and may support the development of more flexible and inclusive strategies for precision immunotherapy in sepsis.

## Declarations

### Author Contribution Statement

ASB contributed to the conception and design of the study. RB and ASB contributed to data acquisition. ASB performed data analysis and performed data interpretation. RB and ASB drafted the manuscript and critically revised the manuscript for important intellectual content. All authors approved the final version of the manuscript and agree to be accountable for all aspects of the work.

### Ethics

This study was conducted in accordance with the principles of the Declaration of Helsinki and was approved by the Institutional Review Board (IRB #15328 and #27044) of the participating academic medical center. For critically ill patients, eligibility screening was performed using electronic medical records and confirmed by clinician review. Given the acute nature of critical illness, informed consent was obtained from patients or their legally authorized representatives, as appropriate.

## Data Availability

All data produced in the present study are available upon reasonable request to the authors

## Acknowledgements

The authors used ChatGPT (OpenAI) to assist with grammatical editing and improvement of manuscript phrasing. The authors reviewed and approved all final content and take full responsibility for the accuracy and interpretation of the manuscript.

---

**Supplemental Figure 1:**
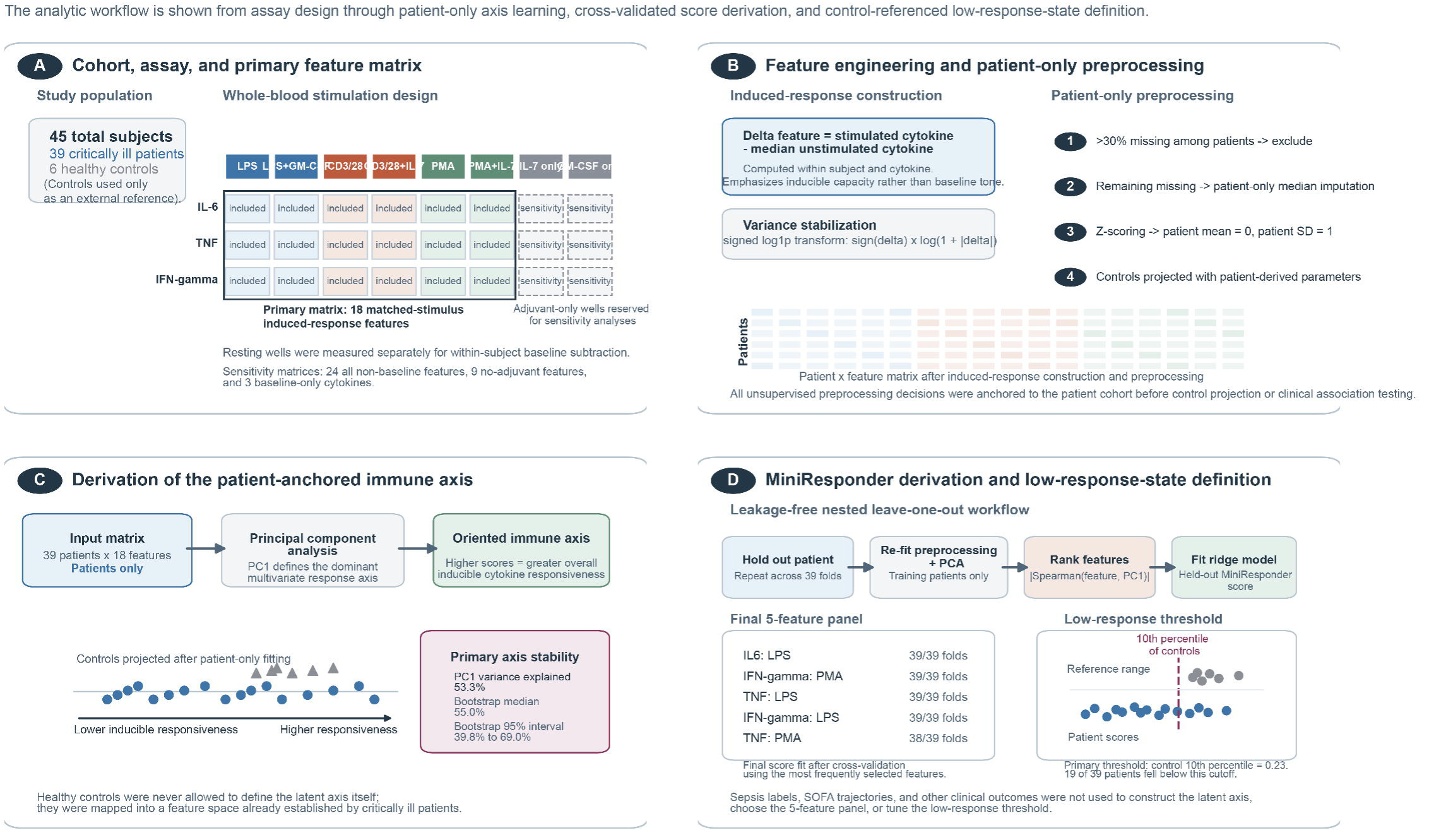
Derivation of the patient-anchored immune axis and MiniResponder score. Panel A summarizes the study cohort and assay structure. Whole blood from critically ill patients and healthy controls was incubated under standardized stimulation conditions, and IL-6, TNF, and IFN-gamma were quantified after incubation. The primary analytic matrix comprised 18 matched-stimulus induced-response features; adjuvant-only wells were reserved for sensitivity analyses. Panel B shows feature engineering and preprocessing. For each subject and cytokine, stimulated values were referenced to that subject’s own unstimulated value to create induced-response features, which were then transformed with a signed log1p transform. Features with more than 30% missingness among patients were excluded, remaining missing values were imputed using patient-only medians, and standardization was performed using patient-only means and standard deviations. Panel C shows patient-anchored immune-axis derivation. Principal component analysis was performed using critically ill patients only, and the first principal component was oriented so that higher values represented greater overall inducible responsiveness. Healthy controls were projected into the resulting feature space after patient-only fitting. Panel D shows derivation of the *MiniResponder* score and the exploratory low-response state. A leakage-free nested leave-one-out workflow repeated preprocessing, principal component analysis, feature ranking, and ridge-regression fitting within each training fold to generate held-out patient scores. The final 5-feature panel was assembled from the most frequently selected features across folds. A low-response state was defined post hoc for enrichment-style analyses as a *MiniResponder* score below the 10th percentile of the healthy-control distribution; clinical outcomes were not used to construct the immune axis, select the panel, or define the threshold.

## References

1. Giamarellos-Bourboulis EJ, Aschenbrenner AC, Bauer M, Bock C, Calandra T, Gat-Viks I, et al. The pathophysiology of sepsis and precision-medicine-based immunotherapy. Nat Immunol. 2024;25(1):19–28.

2. Wiersinga WJ, van der Poll T. Immunopathophysiology of human sepsis. EBioMedicine. 2022;86:104363.

3. Bernard GR, Vincent JL, Laterre PF, LaRosa SP, Dhainaut JF, Lopez-Rodriguez A, et al. Efficacy and safety of recombinant human activated protein C for severe sepsis. N Engl J Med. 2001;344(10):699–709.

4. Ranieri VM, Thompson BT, Barie PS, Dhainaut JF, Douglas IS, Finfer S, et al. Drotrecogin alfa (activated) in adults with septic shock. N Engl J Med. 2012;366(22):2055–64.

5. Venet F, Monneret G. Advances in the understanding and treatment of sepsis-induced immunosuppression. Nat Rev Nephrol. 2018;14(2):121–37.

6. Davenport EE, Burnham KL, Radhakrishnan J, Humburg P, Hutton P, Mills TC, et al. Genomic landscape of the individual host response and outcomes in sepsis: a prospective cohort study. Lancet Respir Med. 2016;4(4):259–71.

7. Scicluna BP, van Vught LA, Zwinderman AH, Wiewel MA, Davenport EE, Burnham KL, et al. Classification of patients with sepsis according to blood genomic endotype: a prospective cohort study. Lancet Resp Med. 2017;5(10):816–26.

8. Sweeney TE, Perumal TM, Henao R, Nichols M, Howrylak JA, Choi AM, et al. A community approach to mortality prediction in sepsis via gene expression analysis. Nat Commun. 2018;9(1):694.

9. Antcliffe DB, Burnham KL, Al-Beidh F, Santhakumaran S, Brett SJ, Hinds CJ, et al. Transcriptomic Signatures in Sepsis and a Differential Response to Steroids. From the VANISH Randomized Trial. Am J Respir Crit Care Med. 2019;199(8):980–6.

10. Stanski NL, Wong HR. Prognostic and predictive enrichment in sepsis. Nat Rev Nephrol. 2020;16(1):20–31.

11. Leventogiannis K, Kyriazopoulou E, Antonakos N, Kotsaki A, Tsangaris I, Markopoulou D, et al. Toward personalized immunotherapy in sepsis: The PROVIDE randomized clinical trial. Cell Rep Med. 2022;3(11):100817.

12. Joshi I, Carney WP, Rock EP. Utility of monocyte HLA-DR and rationale for therapeutic GM-CSF in sepsis immunoparalysis. Front Immunol. 2023;14:1130214.

13. Döcke WD, Randow F, Syrbe U, Krausch D, Asadullah K, Reinke P, et al. Monocyte deactivation in septic patients: restoration by IFN-gamma treatment. Nat Med. 1997;3(6):678–81.

14. Albert-Vega C, Tawfik DM, Trouillet-Assant S, Vachot L, Mallet F, Textoris J. Immune Functional Assays, From Custom to Standardized Tests for Precision Medicine. Front Immunol. 2018;9:2367.

15. Giamarellos-Bourboulis EJ, Kotsaki A, Kotsamidi I, Efthymiou A, Koutsoukou V, Ehler J, et al. Precision Immunotherapy to Improve Sepsis Outcomes: The ImmunoSep Randomized Clinical Trial. JAMA. 2026;335(9):775–86.

16. Yang Y, Zhang Y, Wu J, Liu Y, Lei X. Decoding immune low-response states in sepsis: single-cell and 3D spatial transcriptomic insights into immunoparalysis. Front Immunol. 2025;16:1696914.

17. Arapis A, Panagiotopoulos D, Giamarellos-Bourboulis EJ. Recent advances of precision immunotherapy in sepsis. Burns Trauma. 2025;13:tkaf001.

18. Monneret G, Lepape A, Voirin N, Bohe J, Venet F, Debard AL, et al. Persisting low monocyte human leukocyte antigen-DR expression predicts mortality in septic shock. Intensive Care Med. 2006;32(8):1175–83.

19. Samuelsen AM, Halstead ES, Lehman EB, McKeone DJ, Bonavia AS. Predicting Organ Dysfunction in Septic and p Patients: A Prospective Cohort Study Using Rapid Ex Vivo Immune Profiling. Crit Care Explor. 2024;6(7):e1106.

20. Samuelsen A, Lehman E, Burrows P, Bonavia AS. Time-dependent variation in immunoparalysis biomarkers among patients with sepsis and critical illness. Front Immunol. 2024;15:1498974.

21. Bonavia AS, Samuelsen A, Chroneos ZC, Halstead ES. Comparison of Rapid Cytokine Immunoassays for Functional Immune Phenotyping. Front Immunol. 2022;13:940030.

22. Bonavia AS, Samuelsen A, Liang M, Hanson J, McKeone D, Chroneos ZC, et al. Comparison of whole blood cytokine immunoassays for rapid, functional immune phenotyping in critically ill patients with sepsis. Intensive Care Med Exp. 2023;11(1):70.

23. Singer M, Deutschman CS, Seymour CW, Shankar-Hari M, Annane D, Bauer M, et al. The Third International Consensus Definitions for Sepsis and Septic Shock (Sepsis-3). JAMA. 2016;315(8):801–10.

24. van Amstel RBE, Kennedy JN, Scicluna BP, Bos LDJ, Peters-Sengers H, Butler JM, et al. Uncovering heterogeneity in sepsis: a comparative analysis of subphenotypes. Intensive Care Med. 2023;49(11):1360–9.

25. Seymour CW, Kennedy JN, Wang S, Chang CH, Elliott CF, Xu Z, et al. Derivation, Validation, and Potential Treatment Implications of Novel Clinical Phenotypes for Sepsis. JAMA. 2019;321(20):2003–17.

26. Alevizou A, Soulioti P, Giamarellos-Bourboulis EJ, Safarika A. Biomarker guided immunomodulatory precision medicine to improve prognostic, predictive and adaptive enrichment strategies in sepsis trials. Expert Rev Mol Diagn. 2025;25(7):357–69.

