## Supplementary Methods for "Ex Vivo Immune Profiling Defines a Continuous Functional Immune Axis and a Sepsis-Enriched Low-Response State in Critical Illness"

**Supplementary Material**

**Supplementary Methods**

**Detailed Feature Engineering**

The non-baseline assay matrix contained 24 cytokine-condition features spanning 3 cytokines across 8 non-baseline stimulation/adjuvant combinations (**Supplementary Figure 1**). The primary analysis used an 18-feature matched-stimulus matrix comprising induced responses measured under LPS, CD3/28, and PMA stimulation conditions, with or without the matched adjuvant. Adjuvant-only wells (IL-7 or GM-CSF without a primary stimulant) were excluded from the primary axis because they did not align with the primary stimulus-response hypothesis. Prespecified sensitivity analyses also considered the full 24-feature non-baseline matrix, a 9-feature no-adjuvant matrix, and a 3-feature baseline-only matrix.

For each subject and cytokine, the median unstimulated value was subtracted from stimulated measurements to generate delta-induction features. These deltas were transformed using a signed log1p transformation to stabilize variance while preserving response directionality.

Features with greater than 30% missingness among patients were excluded. Remaining missing values were imputed using patient-only medians, and features were standardized using patient-derived means and standard deviations.

**Cross-Validated *MiniResponder* Derivation**

*MiniResponder* was derived using a nested leave-one-out framework. For each held-out patient, preprocessing, one-component principal component analysis, and feature ranking were repeated using the training patients only. Features were ranked by the absolute Spearman correlation between each feature and the training-fold first principal component. A ridge-regression model was then fit using the 5 highest-ranked features and applied to the held-out patient to generate an out-of-fold *MiniResponder* score.

After cross-validation, a final 5-feature panel was assembled using the features most frequently selected across folds and was applied to all subjects for descriptive analyses and figure generation.

**Axis Stability and Threshold Sensitivity**

Axis stability was summarized by bootstrap resampling of the patient-only primary matrix and by comparing alternative feature sets to the primary axis using Spearman rank correlation and dynamic-range preservation. For the exploratory low-response state, sensitivity analyses examined alternate control-based thresholds, including the 5th and 20th percentiles of controls, mean minus 1 standard deviation, and mean minus 1.5 standard deviations.

**Detailed SOFA Endpoint Definitions**

The primary pilot SOFA endpoint was day 1 SOFA minus the mean SOFA across days 3, 5, 7, and 9. Secondary endpoints included the analogous day 15 mean change, available-case and last-observation-carried-forward versions of these endpoints, a day 1-to-day 9 SOFA slope, and binary achievement of a 1.4-point or greater SOFA improvement threshold.

Baseline-adjusted linear models were used for continuous SOFA-change endpoints, and baseline-adjusted logistic models were used for binary SOFA-improver endpoints, each including day 1 SOFA and *MiniResponder* as predictors. Confidence intervals for the *MiniResponder* term were estimated using percentile bootstrap resampling.
